# Using opioid analgesia for chronic pain in adults aged 85+: a qualitative study

**DOI:** 10.64898/2026.06.08.26354706

**Authors:** Alice Faux-Nightingale, Charlotte Woodcock, Christine Walker, Helen E Smith, Victoria K Welsh

## Abstract

**Background:** Chronic pain is common in adults aged 85 years and older (85+) and is associated with detrimental outcomes. Chronic pain guidelines advise first line management with non-pharmacological measures; paracetamol and non-steroidal anti-inflammatory drugs are the preferred analgesics. Challenges in accessing non-pharmacological therapies for adults aged 85+, and the presence of multimorbidity and polypharmacy, mean that opioid medication is often prescribed for chronic pain despite the potential for opioid-related adverse effects and guidance identifying long-term opioids for chronic pain as a potentially inappropriate prescription.

**Aim:** This study aims to explore patient, caregiver, and healthcare professional perspectives on the prescription of opioid medications for pain management for chronic pain in adults aged 85+ to support development of resources for optimising opioid prescribing.

**Design and Setting:** In this qualitative study, participants were recruited through primary care, in the community or in care home settings.

**Method:** 36 semi-structured interviews were conducted with care home residents and community dwellers aged 85+ (n=12), caregivers (informal and care home staff) (n=12), and healthcare professionals (n=12). Interviews were transcribed and analysed using reflexive thematic analysis.

**Results:** Four themes were developed: contextual complexity, satellite influences, balancing act, and pragmatic prescribing. Using opioids in adults aged 85+ is a balancing act to support patients’ best possible quality of life within their unique circumstances whilst using the pain management tools available.

**Conclusion:** Opioids continue to have an important role in pain management in adults aged 85+ largely due to paucity of alternatives and the drive to support quality of life.

**Key points:**

1. Managing chronic pain in adults aged 85+ is complex; a pragmatic approach to opioids is adopted by patients, caregivers, and prescribers;
2. A focus on improved access to non-pharmacological therapies may offer alternatives to opioids;
3. Emerging Integrated Neighbourhood Teams in England presents a valuable opportunity to support chronic pain for adults aged 85+.

## Introduction

In the UK, the number of adults aged 85 years and older (85+) is predicted to double from 1.7 million in 2022 to 3.3 million in 2047^1^; a trend reflected globally with the number of adults aged 80 years and older expected to triple between 2020 and 2050 to 426 million.^2^ In 2021, there were 278,946 adults aged 65+ living in care homes in England and Wales.^3^

Chronic pain, defined as pain present for more than 12 weeks^4^, is common in people aged 85+. Prevalence estimates of chronic pain in adults aged 85+ range from approximately 50% to 72% in high-income countries, and up to 62% in low- and middle-income countries.^5^ Prevalence of chronic pain in nursing home residents has been estimated to be between 55.9-58.1%.^6^ Causes of chronic pain in this age-group are wide-ranging and include musculoskeletal conditions, neuropathies, and cancer.^7^ The presence of chronic pain in older people is associated with detrimental outcomes including limited mobility and falls,^8^ both of which can lead to loss of independence and reliance on others to assist with activities of daily living.^9^

Guidelines for managing chronic pain recommend a non-pharmacological approach (healthy weight, physical activity, physical therapy, psychological intervention) first (NICE^10,11^, RACGP^12^), with paracetamol and non-steroidal anti-inflammatory drugs (NSAIDs) as the preferred analgesics. The prescription of opioid analgesics (including codeine, tramadol, morphine, buprenorphine, fentanyl) are advised in the lowest dose for the shortest time^13^, or not to initiate at all^14^ due to their addictive properties and subsequent risk of dependence.

There are many difficulties documented about accessing non-pharmacological therapies for the people aged 85+, including physical immobility^15^, beliefs around medication use ^16, 17^, and fear of falling due to pain.^18^ Difficulty engaging in services supporting physical activity can itself lead to further reduction in movement, progression of sarcopenia and resulting increased frailty, and the associated social isolation^19^ and mood disturbance.^20^

Adults aged 85+ are often living with multiple long-term conditions leading to polypharmacy and increasing frailty requiring support from caregivers. The physiological process of ageing is associated with pharmacokinetic and pharmacodynamic changes that increase susceptibility to adverse drug events, including reduction in renal and hepatic clearance of medicines^21^, and increases in drug receptor sensitivity.^22^ For these reasons, NSAIDs are often contraindicated in the people aged 85+, leading to the consideration of opioid medicines when paracetamol is insufficient.

Opioids have many adverse effects including constipation, nausea and vomiting, respiratory depression, drowsiness, confusion^23^, and falls^24^; the physiological changes of ageing increase the risk of these occurring. The increased risk of opioid-induced adverse drug events and the dearth of evidence supporting the use of opioids to manage chronic pain in adults aged 85+ have led deprescribing guidelines to identify long-term opioids for chronic pain as a potentially inappropriate prescription^25^ that clinicians should review and consider stopping. However, opioids continue to be prescribed for chronic pain to adults aged 85+.

To support appropriate opioid prescribing in adults aged 85+, an understanding of the decision-making involved in the prescribing of these medicines from the perspectives of key stakeholders is required including caregivers. There are a myriad of studies investigating beliefs towards prescribing of opioid medicines for chronic pain, though none are designed specifically to address this in the context of adults aged 85+, which is surprising given the physiological, social, and psychological factors that need considering when making pain management decisions, particularly in a population more susceptible to adverse drug events. Our study therefore seeks to bridge this knowledge gap by exploring patient, caregiver, and health care professional perspectives on the prescription of opioid medications for pain management for chronic pain in adults aged 85+.

## Methodology

Ethics approval was granted from the West of Scotland Research Ethics Committee (23/WS/0118).

### Recruitment

Three participant groups were recruited: adults aged 85+ living either in the community or within care homes (patients), caregivers (including relatives), and healthcare professionals (HCPs). Purposive sampling with a focus on maximum variation^26^ was used to recruit a diverse range of participants within each group, sampling on age, ethnicity, gender, location of residence (patients), and professional role (HCPs). Participants were informed about the study and had time to consider and ask questions prior to consent and participation.

Community-dwelling patients were invited to take part in the study, by their general practice, if they were aged 85 years or older, were registered with a general practice within the West Midlands Clinical Research Network (WM CRN), and had a prescription issued for an opioid medicine in the previous three months and one week together with at least one prior prescription in the preceding 12 months.

Patients were excluded if they were in receipt of end-of-life care, if they did not have capacity to consent for themselves, or they were unable to meaningfully communicate about their experiences in an interview. Eligible patients were posted a study information pack with the study information leaflet and consent form. Study team contact details were shared to provide more information or translation. Two care homes were identified separately to ensure the inclusion of patients in care home settings, eligible patients were identified and initially invited by care home staff.

Caregivers were eligible to participate if they were aged 18 years and older, had been identified by the patient as offering support, or self-reported supporting older family members, or were employed in a caregiving role. Caregivers working in care home settings were identified and invited to the study through the WM CRN Enabling Research In Care Homes (ENRICH) team, who distributed study information packs containing a caregiver-specific study information leaflet, consent form, and study team contact details.

HCPs were eligible if they were involved in the care of adults aged 85+ including the prescribing or administering of opioid medicines, or discussion about pain and opioid use. Potentially eligible participants were identified using professional networks and sent study information to inform and invite them to the study.

### Data generation

Interviews took place from January to March 2024, lasting between 10 and 74 minutes (average 32 minutes). Semi-structured interviews were conducted by two experienced qualitative researchers, both female, one non-clinical health services researcher (AFN, MPhil), the other, a general practitioner (GP) (VW, PhD). Patient interviews were conducted in person or by telephone; caregiver interviews were conducted in person, by telephone, or by video call (Microsoft Teams); HCP interviews were conducted in person or by video call, as chosen by the participant. Interviews were offered as one-to-one with the researcher, though where participants requested that their interview took place with other participants (e.g. patient and caregiver, or multiple HCPs), this was permitted. Neither researcher had a relationship with the patient participants prior to data collection. There were instances where HCPs or caregivers were known to at least one member of the research team prior to the interview. Where possible, participants were interviewed by an unfamiliar member of the research team, but if this were not possible the team reflected on the relationship between interviewer and interviewee and the consequences that this may have upon the generated data during analysis.

Topic guides were developed with the study patient and public involvement and engagement (PPIE) group and were used flexibly in interviews to allow the researcher to pursue participants’ comments. Topic guides (Appendix 1) were revised iteratively to reflect and adapt to comments in previous interviews.

All participants provided consent before their interview. Interviews were audio-recorded (in-person and telephone) or video-recorded (video call) with consent. All interviews were transcribed by a professional transcription company and were pseudonymised before analysis.

### Analysis

Interpretive description methodology guided our analysis^27^ utilising clinical knowledge and expertise within the team and acknowledging a practice-orientated focus to the analysis as we developed findings to support future clinical practice. An inductive, reflexive thematic analysis of the interviews was based on the principles outlined by Braun & Clarke.^28^ As part of this analysis, we considered participants’ experiences according to participant group, relationships across the groups e.g. known connections between patients and caregivers, and as a complete dataset to holistically explore the experiences and interactions of the participants. Transcripts were coded inductively by AFN. Themes and subthemes were generated iteratively with the whole interdisciplinary research team, allowing each individual to contribute additional insight and perspectives from their respective disciplines and clinical background. Themes were developed according to commonality across the dataset, significance, and relevance to the research questions. AFN and CWo are qualitative health researchers, CWa is our lay co-author with relevant lived experiences, and VKW and HES are research-active general practitioners with experience of working with older adults who are prescribed opioids for chronic pain. A nurse specialising in geriatric care also supported analysis alongside research question and design, KL. These findings were discussed and refined with our PPIE group who supported development of our key messages and associated patient and caregiver resource.

### Patient and public involvement

Our PPIE group had lived experience of chronic pain and management, are either aged 85+, and/or have experience of caring for older people. This group, alongside our lay colleague, CWa, was involved throughout the study and contributed to the initial design of the research project, public-facing documents, the research findings, and dissemination plans.

## Findings

### Sample characteristics

Forty-nine eligible participants expressed interest in participating, 37 were interviewed (Table 1)

**Table 1:**
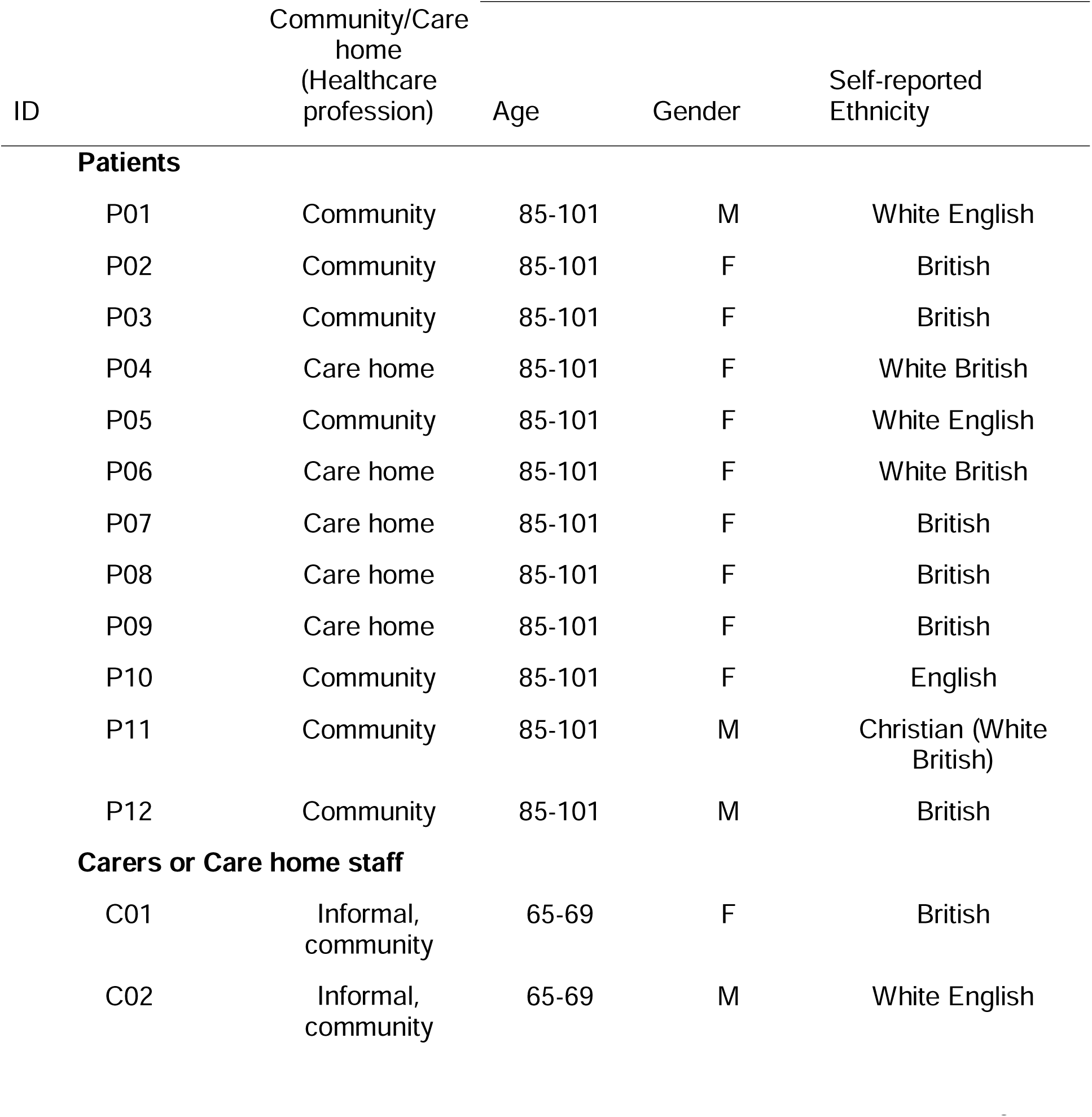
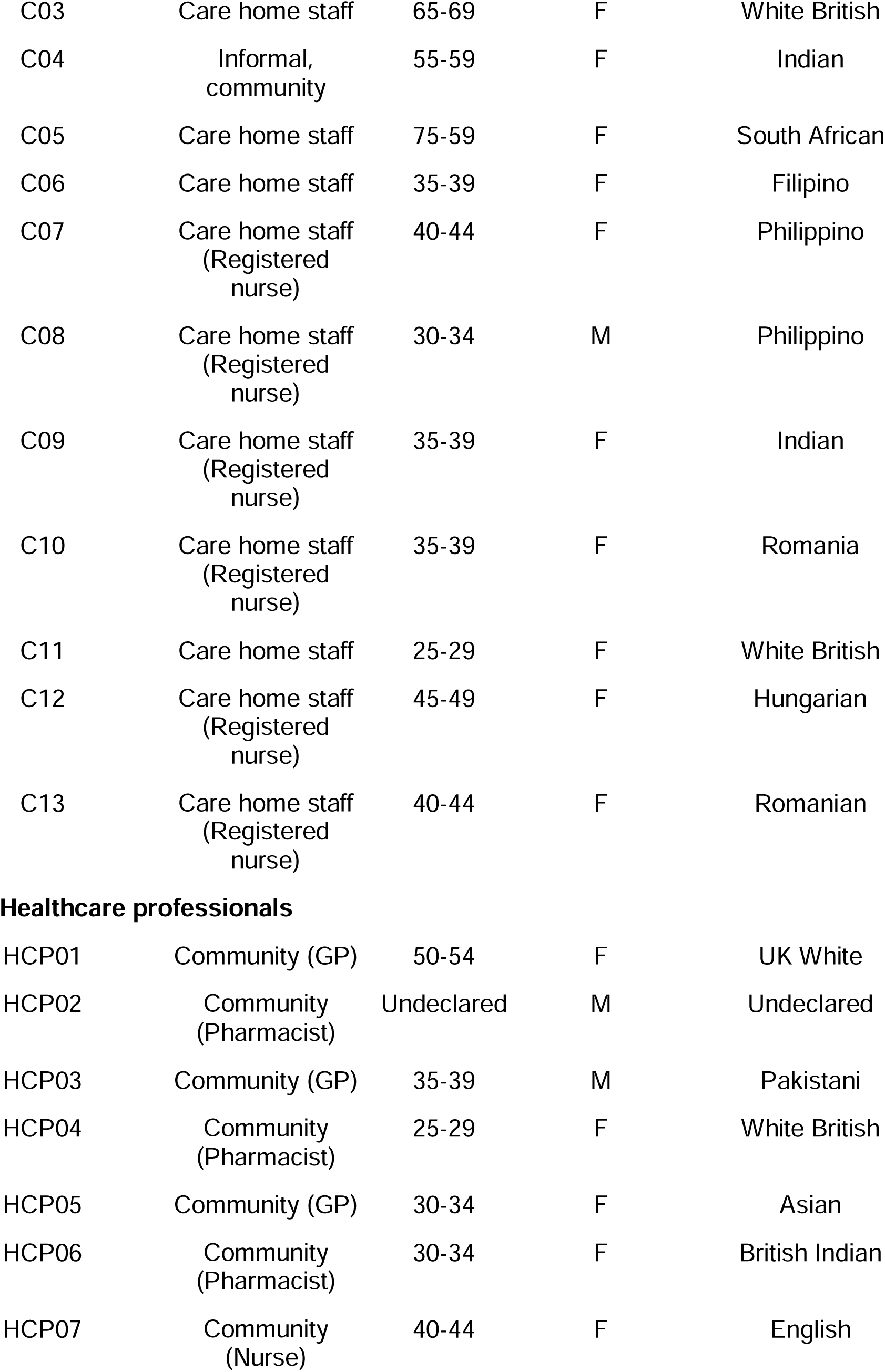
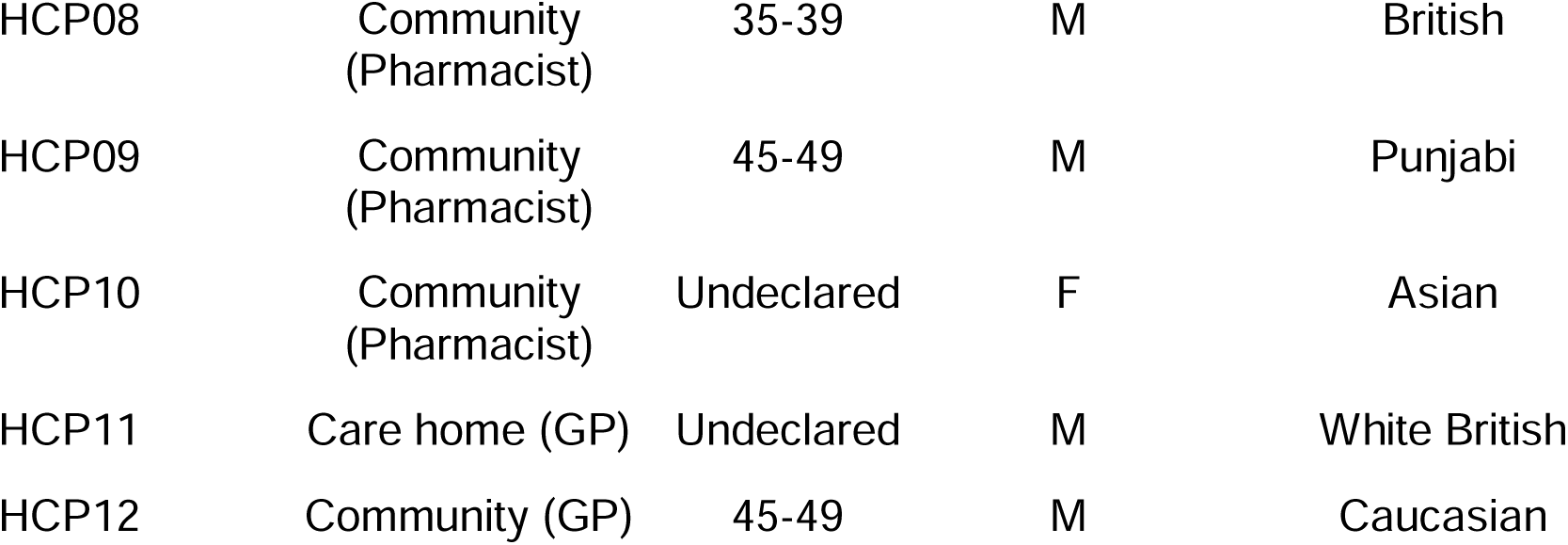
Participant details.

| ID | Community/Care home<br>(Healthcare profession) | Age | Gender | Self-reported Ethnicity |
| --- | --- | --- | --- | --- |
| **Patients** |  |  |  |  |
| P01 | Community | 85-101 | M | White English |
| P02 | Community | 85-101 | F | British |
| P03 | Community | 85-101 | F | British |
| P04 | Care home | 85-101 | F | White British |
| P05 | Community | 85-101 | F | White English |
| P06 | Care home | 85-101 | F | White British |
| P07 | Care home | 85-101 | F | British |
| P08 | Care home | 85-101 | F | British |
| P09 | Care home | 85-101 | F | British |
| P10 | Community | 85-101 | F | English |
| P11 | Community | 85-101 | M | Christian (White British) |
| P12 | Community | 85-101 | M | British |
| **Carers or Care home staff** |  |  |  |  |
| C01 | Informal, community | 65-69 | F | British |
| C02 | Informal, community | 65-69 | M | White English |
| C03 | Care home staff | 65-69 | F | White British |
| C04 | Informal,<br>community | 55-59 | F | Indian |
| C05 | Care home staff | 75-59 | F | South African |
| C06 | Care home staff | 35-39 | F | Filipino |
| C07 | Care home staff<br>(Registered<br>nurse) | 40-44 | F | Philippino |
| C08 | Care home staff<br>(Registered<br>nurse) | 30-34 | M | Philippino |
| C09 | Care home staff<br>(Registered<br>nurse) | 35-39 | F | Indian |
| C10 | Care home staff<br>(Registered<br>nurse) | 35-39 | F | Romania |
| C11 | Care home staff | 25-29 | F | White British |
| C12 | Care home staff<br>(Registered<br>nurse) | 45-49 | F | Hungarian |
| C13 | Care home staff<br>(Registered<br>nurse) | 40-44 | F | Romanian |
| **Healthcare professionals** |  |  |  |  |
| HCP01 | Community (GP) | 50-54 | F | UK White |
| HCP02 | Community<br>(Pharmacist) | Undeclared | M | Undeclared |
| HCP03 | Community (GP) | 35-39 | M | Pakistani |
| HCP04 | Community<br>(Pharmacist) | 25-29 | F | White British |
| HCP05 | Community (GP) | 30-34 | F | Asian |
| HCP06 | Community<br>(Pharmacist) | 30-34 | F | British Indian |
| HCP07 | Community<br>(Nurse) | 40-44 | F | English |
| HCP08 | Community<br>(Pharmacist) | 35-39 | M | British |
| HCP09 | Community<br>(Pharmacist) | 45-49 | M | Punjabi |
| HCP10 | Community<br>(Pharmacist) | Undeclared | F | Asian |
| HCP11 | Care home (GP) | Undeclared | M | White British |
| HCP12 | Community (GP) | 45-49 | M | Caucasian |

### Analysis

Four themes were developed exploring the complexities of the prescription and use of opioid medications for pain management in adults aged 85+: contextual complexity, satellite influences, balancing act, and pragmatic prescribing.

#### Contextual complexity

Participants’ narrative around opioid prescribing and use was grounded in the specific needs and context of adults 85 years and older, including multimorbidity, polypharmacy, stage of life, frailty, and resulting consequences.

> *“Because, as you know, patients don’t just have one morbidity, there’s a whole range, and there’s a pretty good chance that if they have pain, they’ll probably have some other morbidities alongside that as well. So, I think the challenge, for me, is that the older patients fall into that category.” (HCP02, pharmacist)*

Participants described how opioids can reduce pain, support daily activities and improve overall quality of life for those 85 years and older.

> *“I mean, generally, when I review the older patients, I find that they do not want to be taken off these medications, because they feel like it helps with their quality of life. With i.e. managing their pain.” (HCP07, pharmacist)*
>
> *“I think [co-codamol] it’s a brilliant drug. […] it does give people a better quality of life. I don’t think anything else can do the job.” (P17)*

#### Satellite influences

It was common, but not ubiquitous, for community-residing patients to mention satellite figures (e.g. spouse, other relative) who supported them in accessing healthcare and attending appointments and, in some cases, contributed to the decision-making process with respect to opioid prescribing. Residents in care homes had similar support, in addition to that of care home staff.

> *“It’s like everything; haven’t only got to manage the expectations of the patient, but also the family around them, and then the nursing staff around them.” (HCP14, GP)*

Participants across all groups gave examples of how satellite figures could influence prescribing decisions. The impact if this influence varied according to the relationship between the patient and the satellite influencer, and the perceptions and preferences of the satellite influencer. Some caregivers described being present to support: family members navigating their prescription, for example when arranging appointments; with transport; in the appointment itself, acting as an interpreter, advocate, or memory aid; monitoring the impact of new medications and monitoring side effects; and communicating with HCPs as necessary. The caregiver often brought a more holistic view of medication impact and supported the older person assessing the risk-benefit of opioid side effects, all factors which supported the HCP in making (and revisiting) prescribing decisions.

> *“[I] did say to her, ‘Is it taking the pain away?’ And she said no. So, you’re getting no benefit and you just get sick really?” (C04)*

However, these satellite influences could also be disadvantageous; a third party with strong views that differ from those of the patient on whether their relative should or should not take opioid medications, could steer the decision-making away from the patient’s preference.

> *“It depends on the patient, and it depends on who else is in the room, because in that age group, they’ve often had someone bring them to the appointment. You may have daughter or son in the room saying, ‘You’re in pain, we need to get rid of this pain for you, mom/dad.’ And the patient is not really wanting to take anything.” (HCP01, GP)*

#### Balancing act

Prescribing opioid medications to older people appeared to be a careful decision-making process which balanced the benefit of pain relief for the patient, in terms of symptoms and quality of life, with the side effects and risk of adverse effects of opioids. This process continually adjusted as patients, caregivers, and HCPs monitored for medication efficacy, adverse effects, and long-term tolerance; all considered on an individual level to meet the needs and circumstances of the patient.

> *“They are all very individual patients and there are many factors in what people think may help with their pain.” (HCP03, GP)*

For HCPs, the main aim in prescribing opioid medications was to reduce patients’ pain, and in doing so, improve overall quality of life. Age, particularly the proximity to the end of life, also factored into these considerations:

> *“[the patients say] ’at my age, if I can get a bit of pain relief, I’m quite happy to take opioids.’ Because it’s about quality of life, isn’t it?” (HCP02, pharmacist)*

While all participants perceived the need for balance, different groups approached consideration of pros and cons differently. For patients and family caregivers, the perceived benefit of pain relief often out-weighed the con of adverse effects, that were tolerated and deemed easy to manage at home.

> *“If it does the job, great, I’ll take it, you know. Whatever, as long as it does the job.” (P03)*

HCPs, however, gave more weight to the risks, discussing the potential for adverse drug reactions and the interactions with other medications, weighing these against patients’ needs and social arrangements. There was some concern that older people do not consider these risks appropriately:

> *“but often the elderly, they do underestimate, you know, potentially the risks of some of the medications,” (HCP03, GP)*

The decision to prescribe opioid medications to older people was often described as the least worst option available, an imperfect balance of benefit to risk:

> *“Rarely people just come in just because they have an occasional pain […] They’re coming because they cannot walk, they cannot do usual things okay so it’s almost finding the […] lesser evil” (HCP15, GP)*

This balancing act, however, was seemingly done on an individual basis both in terms of individual clinician behaviour and individual patient factors. Individual differences may be influenced by the longevity of clinical practice, experience of opioid prescribing in the adults aged 85+ and utilisation of guidelines to support their decision-making. For example, one experienced pharmacist referred to the dearth of nuanced guidelines for chronic pain in adults aged 85+ as a possible reason for weighting the decision towards risks and thus not prescribing opioids in this situation:

> *“It’s probably good to have a resource around just managing opioids in this group, as we do with children […] in terms of how weight loss, or BMI, or your other organs that function, how it does affect [them]? What I do find with some of the younger pharmacists, I think they sometimes want the decisions making for them. So, they do [use] the pathways and flowcharts.” (HCP11, pharmacist)*

#### Pragmatic prescribing

In addition to the balance between benefit and risk, HCPs also described how the prescription of opioid medications was, in many cases, a difficult but necessary option for this age group; a pragmatic approach to help the individual with their pain. Despite guidelines recommending alternative pain relief options and the deprescription of opioid medications, HCPs, particularly GPs, described the prescribing of opioids necessary because alternative options were not available, due to physical accessibility by the patient, pharmacological intolerance or contraindication, or local availability.

> *“The guidelines say not to use analgesia as the first line, but maybe offer TENS and acupuncture and those… It’s not available. So it’s I think it’s easier sometimes [to prescribe opioid medications].” (HCP11 - pharmacist)*
>
> *“So it’s really limited to what we can say because these people, they’re elderly, they’re frail, how much are you going to say exercise? It’s just this is quite difficult for them. So, it is difficult to suggest an alternative apart from paracetamol, really.” (HCP07, pharmacist)*

There was also acknowledgement of how opioid medications can not only improve pain but subsequently enable access to non-pharmacological methods of pain management.

> *“But if your aim is actually to get up and about, and you know the pain is the barrier, and that’s my conversation. Look, this is. I’m just giving you a different walking stick. […] nobody’s gonna move until somebody does something about the pain.” (HCP15, GP)*

## Discussion

### Summary

Our study explored patient, caregiver, and HCP perspectives on the use of opioid medicines for pain management in adults aged 85+, and developed four overarching themes driving decision-making: the contextual complexity of this age-group with frequent multimorbidity, polypharmacy and frailty; the satellite influences around the patient; the balancing act of prescribing; and pragmatic prescribing as the only available option. Managing pain in this age group is a balancing act carried out between multiple parties, considering the patient’s personal circumstances and risks, the perspectives of the people around them, and the availability of healthcare resources.

### Comparison with existing literature

Research exploring problematic prescribing and prescribing cascades within older people found prescribers were going through similar decision-making processes in relation to prescribing for chronic diseases whereby the complexity of prescribing decisions in the context of polypharmacy made finding the balance between the risk and benefits challenging.^29^ Consequently, HCPs avoided making medication changes unless there was clear evidence of harm, creating what the authors termed a ‘deprescribing inertia’^29^. This mirrors our findings in the struggles that HCPs have in finding that balance and thus deciding to prescribe, particularly with a lack of alternative options. Our study builds on these findings and echoes those from a US-based study exploring the use of opioids for chronic pain in older people^30^ in describing a driver for prescribing opioids for chronic pain as quality of life i.e. a degree of relief from the pain, and a decision to avoid starting opioids, or stopping them, may be even more difficult where the symptom burden is obvious and distressing for all stakeholders. The concept of deprescribing inertia is important for opioid prescribing because, if prescribers are tipping the scales towards starting opioids as a pragmatic choice for pain management in the absence of other strategies, or a perceived greater risk of adverse drug events with alternatives, then it may be that prescriptions are continued due to this sense of deprescribing inertia on behalf of prescribers, thus starting a new potential prescribing cascade.

The British Geriatric Society’s 2025^31^ guidance on pragmatic prescribing for people living with moderate or severe frailty advocates for shared decision-making and more lenient targets that may better balance medication benefits and harms in the management of heart failure, hypertension, and diabetes. An expert commentary points out that pragmatic prescribing is not *a “retreat from evidence-based practice, but instead recommends that evidence be interpreted in the light of lived vulnerability and patient preferences”*^32^, and it appears that the HCPs in our study are actively applying this principle and would welcome clear pragmatic guidance in relation to the management of chronic pain in the context of frailty, polypharmacy, and multimorbidity to help with their decision-making and balancing of benefits and risks.

European recommendations on the pharmacological management of pain in older people have been published since our interviews and go some way towards providing pragmatic guidance on the use of opioids for chronic pain in older people. Derived by a multidisciplinary group of European pain experts,^33^ these advise that, in combination with non-pharmacological approaches, weak opioids have their place in the treatment of chronic pain of moderate-to-severe intensity in older people, starting with low doses with careful monitoring for therapeutic effect and adverse drug reactions. These recommendations also advocate for the involvement of the patient and their family in their treatment echoing our study findings that caregivers can provide valuable insights into adverse drug events and opioid efficacy.^34^

Indeed, caregivers have a critical role to play in medication management, both for the practical aspects obtaining and administering medication, and in the more complex tasks of organising and tracking medication, gathering information, and making treatment decisions^35^; this was evident in our study, with our community caregivers in particular undertaking these jobs. Our HCPs acknowledged the input of caregivers, in keeping with shared decision-making guidelines.^36^ Measures to optimise pain management in adults aged 85+ need to therefore consider the role of caregivers.

In addition to the key drivers highlighted in our study, other influences on opioid prescribing for chronic pain in older adults have been identified in a scoping review,^37^ for example, subjective judgements by prescribers on ‘tolerable pain’^38^, and administrative burden of prescribing.^39^ These topics were not discussed in our interviews, likely due to these findings identified in studies set outside the UK and not within primary care.

### Strengths and Limitations

This study has many strengths. It addresses a paucity in research in this area and offers insight into the experiences and perceptions of key stakeholders involved in the prescription of opioid medications to people aged 85+, including presenting interviews from seldom-heard populations: people aged 85+, and care-home residents and staff. Recruiting from these three populations also enabled us to gain a broader understanding of this area, allowing us to consider the interactions and nuances which underlie any opioid prescriptions to older people.

Whilst we succeeded in recruiting 12 people aged over 85 to participate in this study, these patient participants had limited demographic diversity, all being white-British. Many of our participants outside of the care homes lived independently or with family in their homes, with only one living in assisted living. We acknowledge that there may be other perspectives and experiences we have not captured here and that there may be additional considerations in the prescription process for different settings. Further research in different localities and from different cultural backgrounds would expand our understanding of the prescription process and consider any additional factors in inequities, including within health services, which may affect the prescription process or identify additional needs which should be considered in future guidelines.

Whilst we recruited good numbers of care home staff and health care professionals, we also had limited numbers of family carers, and no domiciliary carers. Efforts to boost recruitment of family carers and get representation from domiciliary carers through advertising on social media and in specific online groups for carers, and approaching carers’ networks yielded no success, potentially due to the intensity and duration of caring roles. Future research would benefit from exploring the experiences of these missing voices.

### Implications for research and practice

Managing chronic pain in adults aged 85 years and older is challenging due to varying levels of co-existing multimorbidity, polypharmacy, and the presence of frailty which preclude non-opioid analgesics and limit access to existing non-pharmacological measures. Shared decision-making is crucial to help weigh the risks and benefits of opioid medication and caregivers have a vital role to play in the monitoring for adverse effects and pain efficacy. Although specific evidence of benefit for behavioural interventions on chronic pain outcomes remains limited^40^, focussing on improving access to non-pharmacological therapies for older adults including physical activity, psychological and psychosocial interventions may offer an alternative to opioid prescribing and thus reduce the risk of adverse effects and resulting prescribing cascades in addition to more general benefits to health that come from these non-pharmacological strategies. The developing Integrated Neighbourhoods Team approach within England as part of the Government’s 10 Year Plan for England^41^ offers the opportunity for developing services designed to support the needs of adults aged 85+ by connecting those accessing health or social care to support from additional sectors including voluntary sector organisations, public health, and local government services within the local community.^42^

As guidelines advise, providing regular medication reviews for patients and, where appropriate, caregivers, will afford the time and space to review pain management and ongoing opioid requirements as the needs, circumstances, and priorities of individuals change. This will also capture patients who have had opioid prescriptions initiated in secondary care. Alongside medication reviews currently in existence for opioids as a ‘high risk medicine’^43^, there may be opportunities to review pain management during proactive approaches to care, for example within a Comprehensive Geriatric Assessment^44^ when triggered by frailty case-finding, or reactively during an assessment for a specific symptom.

Caregivers in our study demonstrated their important role as satellite influencers of the decision-making process relating to using opioids for chronic pain. A systemic review addressing family caregiver challenges in pain management for patients with advanced illnesses found themes of care-giver related fears, beliefs, and function; knowledge and skills in pain management; and communication with HCPs.^45^ The dearth of evidence in non-advanced illness, for example multimorbidity and non-cancer diagnoses was highlighted.^46^ Understanding the needs of caregivers in supporting those in chronic pain through further research, and developing evidence-based caregiver-specific resources focussed on helping to manage chronic pain in older adults with complex health and social care needs, may further optimise the use of opioids in adults aged 85 years and older.

## Conclusion

Prescribing and using opioids in adults aged 85 years and older is a balancing act which looks to support a patient’s best possible quality of life within their unique circumstances, using the pain management tools available. Although there is a general steer towards deprescribing of opioids, in this age-group it appears opioids continue to have an important role in pain management due to lack of other options; an imperfect solution to a difficult situation.

## Data Availability

The datasets generated during and/or analysed during the qualitative work packages will be available upon request from Alice Faux-Nightingale. Any subsequent requests for access to the data from anyone outside of the research team (e.g. collaboration, joint publication, data sharing requests from publishers) will follow the Keele University SOP data sharing procedure. 

## Data sharing

The datasets generated during and/or analysed during the qualitative work packages will be available upon request from Alice Faux-Nightingale. Any subsequent requests for access to the data from anyone outside of the research team (e.g. collaboration, joint publication, data sharing requests from publishers) will follow the Keele University SOP data sharing procedure. <u>Data Management and Access Plan</u>

