## Supplementary material 1 for "Using opioid analgesia for chronic pain in adults aged 85+: a qualitative study"

**Appendix 1: Topic guides**

**Interview topic guide: Patient participants**

A) Can you please tell me about the reasons you started your opioid pain medicine?  
Experience of pain prompts:

- Where was the pain?
- How long had you had the pain?
- Had you tried other medication beforehand?
- How has the pain experience changed over time?
- Do you think that your age affects the way you cope with the pain?
- Does the pain affect the way you relate to people?
- Does the pain affect your everyday life?

B)  Please can you tell me about your experiences of taking your opioid pain medication?:  
Experience of medication prompts:

- How long have you been taking opioid pain medication?
- Have you taken opioid pain medication consistently during this time or has your use varied?
- Can you tell us about the time opioids were started? Were you involved in the decision?
- Do you find them helpful?
- Do you have any side effects?
- Do you have any concerns about taking them long term?
- Do you have any concerns about them being reduced or stopped?
- Do you adjust your pain medication?
- Do you think that your age affects your attitude to the pain medication?
- Could you tell us about anything other than medication that helps with the pain? (Distraction? hot water bottles?)
- Has your ability to access other strategies to control pain changed with time?

C) Please can you tell me about your experiences of talking to the prescriber about taking your opioid medication? 
Talking to healthcare professionals prompts:

- Who do you talk with about your pain medications?
- Are there any problems talking about your pain medication with your doctor or other healthcare professionals?
- Where else have you got information about your pain medication?
- What would support you in discussing your pain medication with your healthcare team/family?
- Do you think that your age affects the way you talk to healthcare professionals, or that they talk to you?

Is there anything else that you want to tell me about your experience with opioid medications or discussing with healthcare professionals?

**Indicative Interview topic guide: Family Carer participants**

A) Experience of pain: Can you please tell us about your understanding of the pain that X has experienced? 
Prompts:

- How do you think X’s pain has changed with time? Now and previously
- How do you think that X’s age has affected the way that they cope with the pain?
- How does X’s pain affect the time that you spent together?
- Do you think that X’s pain has affected your relationship? If so, how?

B) Experience of medication: Please can you tell me what you know about the pain medication that X takes?
Prompts:

- Do you help X with their pain medication?
- If so, has this caused any issues for you?
- Do you talk with X about their pain medication?
- How do you feel about X taking strong pain killers long-term?
- If concerns are raised, have you discussed these with X or anyone else?
- Do you think that X’s age makes any difference to whether strong pain killers are appropriate?

C) Talking with Healthcare professionals: Do you accompany X to healthcare appointments or advocate with them on X’s behalf? Prompts:

- Have you experienced any problems talking to the healthcare professionals who look after X about their pain medication?
- Where else have you got information about pain medication?
- What would support you in discussing X’s pain medication with their healthcare team/family?
- Do you think that their age affects the way you talk to healthcare professionals?

Is there anything else about your experience of supporting X that you want to tell us that about X’s pain medication or talking to healthcare professionals?

**Interview proforma – Health care professionals**

A) General Experience of prescribing: Please can you tell us about your experiences of prescribing opioid medication to the oldest old (85+)?

- What factors are important to you when assessing the pain of the oldest old (85+)
- What factors do you think it is important to consider when prescribing opioid medication to the oldest old with chronic conditions (non-palliative scenarios)?
- Have changes to prescribing guidance regarding opioid medication affected the way that you prescribe opioid medication to the oldest old?
- What has been your experience of tapering opioid medications with the oldest old?
- Have there been any specific scenarios that have affected the way that you approach the prescribing of opioids to the oldest old
- How do you discuss the risks and benefits of opioids with the oldest old age group?
- How do you discuss the risks and benefits of opioids with family carers of the oldest old?
- How do your patients respond when you prescribe them opioid medication?
- Do patients ask for opioid medication?
- Are they reluctant to take opioid medication or do they have any concerns about taking them?
- Do any patients talk about to you about (potential) addiction to opioid medications?

B) Discussion of specific patient (if relevant)

- What were the key factors that you considered when prescribing opioids to X?
